# Quantifying meaningful usage of a SARS-CoV-2 exposure notification app on the campus of the University of Arizona

**DOI:** 10.1101/2021.02.02.21251022

**Authors:** Joanna Masel, Alexandra Shilen, Bruce Helming, Jenna Rutschman, Gary Windham, Kristen Pogreba-Brown, Kacey Ernst

## Abstract

**Objective:** To measure meaningful, local exposure notification usage without in-app analytics.

**Methods:** We surveyed app usage via case investigation interviews at the University of Arizona, with a focus on the period from September 9 to November 28, 2020, after automating the issuance of secure codes to verify positive test results. As independent validation, we compared the number of verification codes issued to the number of local cases.

**Results:** Forty six percent (286/628) of infected persons interviewed by university case investigators reported having the app, and 55% (157/286) of these app users shared their positive SARS-CoV-2 test result in the app prior to the case investigation interview, comprising 25% (157/628) of those interviewed. This is corroborated by a 33% (565/1,713) ratio of code issuance (inflated by some unclaimed codes) to cases. Combining the 25% probability that those who test positive rapidly share their test result with a 46% probability that a person they infected can receive exposure notifications, an estimated 11.4% of transmission pairs exhibit meaningful app usage. High usage was achieved without the use of “push” notifications, in the context of a marketing campaign that leveraged social influencers.

**Conclusions:** Usage can be assessed, without in-app analytics, within a defined local community such as a college campus rather than an entire jurisdiction. With marketing, high uptake in dense social networks like universities makes exposure notification an impactful complement to traditional contact tracing. Integrating verification code delivery into patient results portals was successful in making the exposure notification process rapid.

**3 question summary box:** *What is the current understanding of this subject?:* The extent to which exposure notification technology reduces SARS-CoV-2 transmission depends on usage among infected persons.

*What does this report add to the literature?:* A novel metric estimates meaningful usage, and demonstrates potential transmission reduction on a college campus. Clear benefit was seen from simplifying verification of positive test results with automation.

*What are the implications for public health practice?:* Defined communities can benefit from local deployment and marketing even in the absence of statewide deployment. Lifting current restrictions on deployment would allow more entities such as campuses to copy the model shown here to be successful.

## Introduction

Smartphone applications (apps) can automatically and privately notify individuals of exposure to a person known to be infected with SARS-CoV-2. This technology has the potential, given sufficient uptake, to significantly reduce the spread of SARS-CoV-2.^1^ It does so not by replacing contact tracing, but by making contact notification faster, more scalable, potentially more acceptable due to greater privacy, and more comprehensive by notifying contacts who the person who tested positive does not know personally.^1^ The University of Arizona community piloted the Covid Watch app, which through the Google/Apple Exposure Notification Application Programming Interface (API), uses Bluetooth to measure date, distance, and duration of contact (between phones, as a proxy for contact between their owners). The University of Arizona pilot was made possible when the Arizona Department of Health Services agreed to act as the publisher of the app, and to affirm to Apple and Google their understanding that no app other than Covid Watch could use the same API in Arizona. App users who tested positive for SARS-CoV-2 could upload cryptographic keys that identify Bluetooth tokens that they had previously broadcast locally during their infectious period. Keys were uploaded to a public server that was checked by the apps of other Covid Watch users. Detection of a match corresponding to an above-threshold risk of infection^2^ anonymously triggered a notification, including testing and quarantine recommendations.

The focus of this study is to assess meaningful app uptake within a campus community. App download numbers are readily accessible, but overstate how many apps actively check the server.^3^ What is more, in our case, downloads during our 2020 pilot represent some unknown combination of persons present on the University of Arizona campus, persons present on the Northern Arizona University campus (which also piloted the app), and persons physically present at neither. Cumulative downloads rose from 8,648 on August 23 to 20,392 on September 9 to 51,267 on November 28, clearly exceeding 100% of the population presumed to be physically present on the University of Arizona campus, given that most instruction was online during the Fall 2020 semester.

The potential for exposure notification to prevent SARS-CoV-2 transmission in any case depends not on overall adoption in a community, but on usage specifically among persons who go on to become infected. One concern is that individuals who are more likely to download an exposure notification app are generally more likely to follow public health guidelines, and so may be less likely to be infected by SARS-CoV-2, making population statistics overestimates of effective usage. Effective usage requires not just app installation by both members of a transmission pair, but also reporting by the initial (or primary) infected person of a positive SARS-CoV-2 test result. To prevent malicious use, infected persons are required to prove that they tested positive, by obtaining and entering a secure numeric verification code, which we distributed both via our test results patient portal and via our Campus Health clinic. With the help of a third party, the University of Arizona used social marketing tools to promote app adoption, with a focus on reaching students via identified influencers;^4, 5^ here we assess the outcome using a new metric of app usage.

## Methods

### Data sources

Evaluating apps that use the Google/Apple Exposure Notification API is challenging because of their strict privacy protections;^6^ all exposure information is present only on the phone of the notified person, and cannot be linked to the source of exposure. To perform the evaluation outside the app itself, we therefore incorporated two questions into case investigation interviews: (i) Have you downloaded the Covid Watch app?; (ii) If yes, did you already enter the verification code following your positive test? A portion of the faculty, staff, and students who tested positive through the on-campus testing program were assigned to a university contact tracing team, Student Aid for Field Epidemiology Response (SAFER), by the county health department.^7^ Assignment of cases to the University of Arizona SAFER team varied over the time period covered in this report due to changing patterns of incidence. Cases with on-campus addresses (dorms and Greek housing) were consistently assigned to SAFER throughout this time period. For off-campus cases tested through the University of Arizona testing program, a surge in cases associated with campus necessitated additional support from county investigators, so from September 10 to October 30, 2020, county health department staff investigated all off-campus cases. Beginning October 31, SAFER investigated all cases tested through the University of Arizona testing program and anyone who provided an on-campus address, even if they were tested off-site. The team filtered the case data by known on-campus addresses each day to create these investigation assignments.

Our data span August 23 to November 28, 2020 (Fall semester), during which there was a significant outbreak of COVID-19 in the student population (Figure 1). Upon first launch on August 23, 2020, verification codes, which allow those who have tested positive for SARS-CoV-2 to prove this fact to their app, were given over the phone by Campus Health. Beginning September 9, end users, when viewing their positive test result in the “Test All Test Smart” portal that supports the university’s high-volume diagnostic testing, were prompted to retrieve a 6-8 digit numeric code automatically if they had the Covid Watch app. To evaluate the impact of automated code delivery, we compare data before versus after September 9.

**Figure 1.**
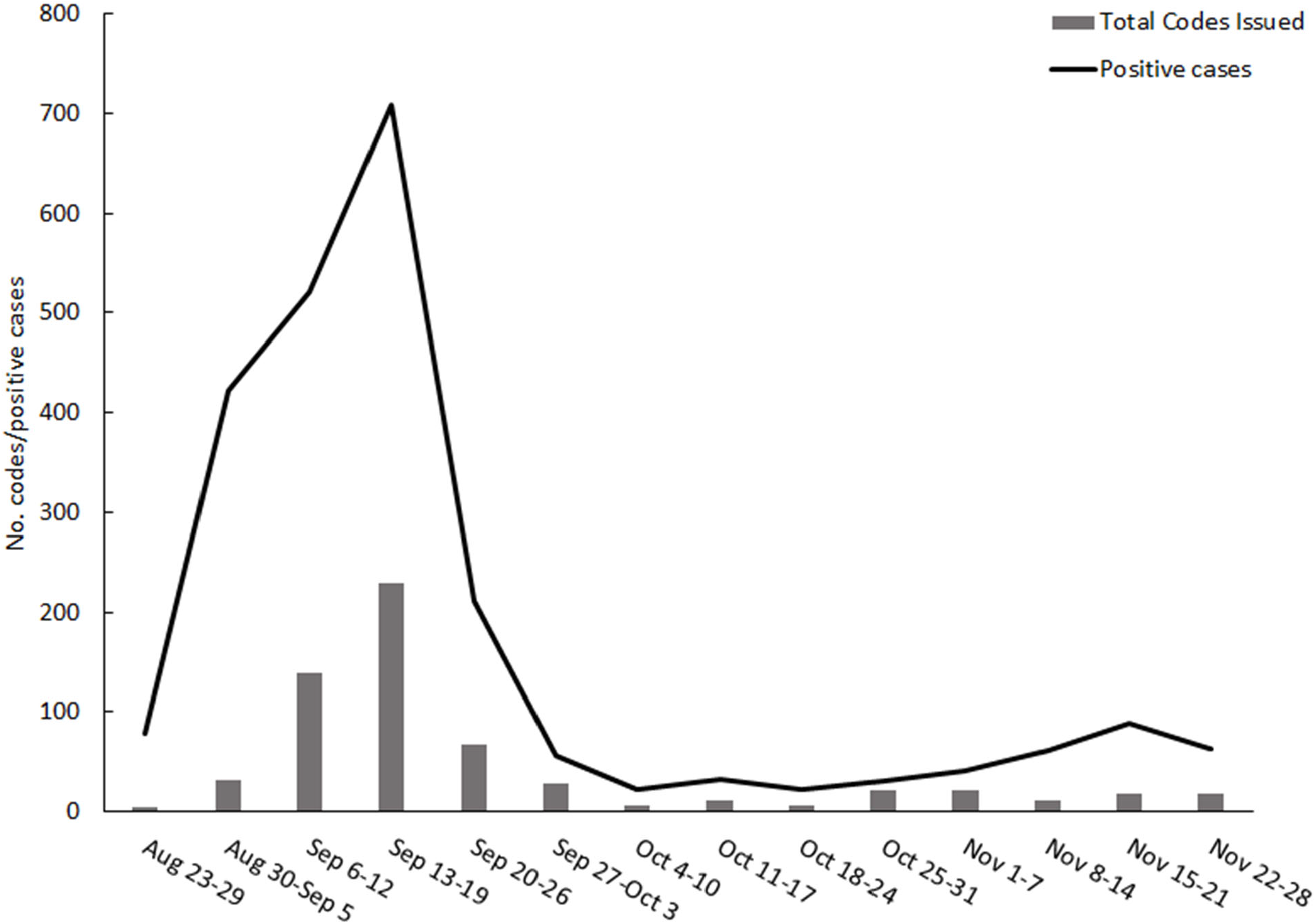
Total verification codes issued (bars) and University of Arizona case counts (line) during Fall semester, 2020. Codes allow app users to prove their positive SARS-CoV-2 test status and thus trigger anonymous notifications in the apps of phones that were previously nearby. Codes were at first delivered over the phone by Campus Health, then beginning September 9 they were also automatically available in the portal with which test results were delivered. The total number of codes issued represented only 7% of the number of cases prior to the automation of code delivery on September 9, and rose to 33% for the remainder of the study period. A reduction in cases was seen shortly after code automation began on September 9, but there is insufficient evidence to causally attribute this to the app, given other control measures put in place around the same time.

Some individuals tested positive twice, e.g. once by antigen test and once by PCR. Positive test results, combined across both Campus Health and the Test All Test Smart program, were de-duplicated to calculate number of cases, by discounting subsequent positive test results for the same person if they occurred within a 90 day window from the first positive test.

Those who agreed to interview may be more likely to follow public health measures, such as app use, than non-interviewed persons with reported positive test results, possibly biasing results towards higher app use. Therefore, we calculated a second metric, the ratio of codes issued / cases. This entailed calculating the fraction of positive tests that led to a request for a verification code by dividing number of codes issued by number of cases. We counted verification codes at point of issue rather than upon usage, but because obtaining a code required action from a person who tested positive (either a phone call or clicking on a request link), we expect this to be only a slight overestimate.

### App marketing

The University of Arizona’s social media-focused marketing campaign surrounding the app launch occurred under the auspices of a pilot in the absence of a State-wide launch. The University website, main app information page, and FAQ page were used as the central source of information, to which all messaging was directed. We used popular University of Arizona hashtags such as #beardown to identify a set of social influencers who we could confirm were enrolled students, and had an Instagram account with more than 4,000 followers. Influencers were given hashtags and example content to share but had full control over their own messages. Example messaging was designed to target individual personas, specifically those who were on the fence about downloading the app. Privacy protection was a key theme to assuage concerns. Influencers were not paid nor given additional incentives such as University apparel. From 978 identified influencers, 148 posted twice or more and an additional 103 posted once within the initial two week launch marketing blitz. Ninety eight identified influencers were student athletes who were unable to post on account of National Collegiate Athletic Association rules and regulations regarding personal influence.

## Results

Campus testing programs recorded 2,728 positive tests from August 23 to November 28, representing 2,360 cases. Of these cases, the University of Arizona SAFER team was assigned 1,359 cases to investigate and conduct contact tracing, while the remaining cases were investigated by the local health department and are not included in our sample (see Methods). The university contact tracing team succeeded in reaching and conducting interviews for 64% (876/1359) of their assigned cases, including the survey questions. Results are summarized in Table 1.

**Table 1.** Key numbers pertaining to app usage by infected individuals at the University of Arizona during Fallsemester, 2020. Codes allow app users to prove their positive SARS-CoV-2 test status and thus trigger anonymous notifications in the apps of phones that were previously nearby. Codes were at first delivered over the phone by Campus Health, then beginning September 9 they were also automatically available in the portal with which test results were delivered. The frequency with which cases used the Covid Watch app increased far more following the automation of verification code delivery than did the frequency with which cases had the app downloaded at the time of case interview. Key numbers following code delivery automation are shown in bold. Cells are left blank when data broken down by date is not available. Positive test results were de-duplicated to calculate number of cases, by discounting subsequent positive test results for the same person if they occurred within a 90 day window from the first positive test.

| **Metric** | **Aug 23 - Sep 8** | **Sep 9 - Nov 28** | **Total** |
| --- | --- | --- | --- |
| Codes issued by phone | 48 | 166 | 214 |
| Codes issued via test results portal | 0 | 399 | 399 |
| Codes issued (total) | 48 | 565 | 613 |
| Positive tests | 676 | 2052 | 2728 |
| Cases | 647 | 1,713 | 2,360 |
| Cases investigated by university contact tracing team |  |  | 1,359 |
| Cases reached by university contact tracing team | 248 | 628 | 876 |
| Downloaded the app | 95 | 286 | 381 |
| Code entered to share their positive test | 32 | 157 | 189 |
| Downloads/Cases reached | 38.3% | **45.5%** | 43.5% |
| Code entries/Cases reached | 12.9% | **25.0%** | 21.6% |
| Codes issued/Cases | 7.4% | **33.0%** | 26.0% |
| Usage metric (product of Downloads/Cases to represent secondary case and Code entries/Cases to represent primary case) | 4.9% | **11.4%** | 9.4% |

The proportion of reported infected persons interviewed who had downloaded the app improved during the initial weeks of the launch, rising from 38% (95/248) before verification code delivery was automated beginning September 9, to 46% (286/628) over the remainder of our reporting period. More strikingly, the proportion of infected, interviewed, and app-using persons who had proved their status as positive for SARS-CoV-2 by entering a verification code into their app prior to the case interview, enabling rapid contact notification, rose from 32/95 (34%) before September 9 to 157/286 (55%) after.

Among those interviewed, 35% (302/876) reported no contacts to case investigators. App usage was similar in this group who reported no contacts: 38.4% (116/302) had downloaded the Covid Watch app, 52.6% (61/116) of whom had entered their code into the app.

From September 9 onward, 25% (157/628) of the interviewed infected persons both had the app and had entered a code to verify a positive test result. Our alternative metric, namely codes issued / cases, rose from 48/647 (7%) prior to September 9 when codes were issued only by phone, to 33% (565/1,713) from September 9 onward.

## Discussion

We propose and estimate a metric of meaningful usage among infected persons. Because the app’s epidemiological impact depends on changing the behavior of infected persons prior to diagnosis, what matters is usage among infected persons, not adoption among the general population. We consider transmission pairs within a tightly interconnected college campus, and estimate the probability that both infected persons have used the app to the minimum necessary level to potentially impact transmission. From September 9 onward, the estimated probability of sufficient usage by a primary, reported infected person is 25% (where verification code entry is required) and 46% for the person they go on to infect (requiring only app activation). Combining these by assuming a population in which any combination of two individuals is equally likely to come into contact, and neglecting transmission from outside campus given low community prevalence at the time of this pilot study, app usage is estimated to affect 11.4% of transmission pairs (Table 1). In a population where individuals in the same transmission pair are more similar than two randomly chosen individuals, including with respect to their propensity to use the app, this value will be higher than 11.4%. If some individuals who infect others never tested positive for SARS-CoV-2, this value will be lower.

With respect to this last caveat, we note that individuals on campus were instructed to test through the university’s “Test All Test Smart” if asymptomatic (which returned 1,841 positive tests), and at Campus Health if symptomatic (which returned 887 positive tests). This is consistent with a high rate of detecting infection, including asymptomatic and presymptomatic infections.

Our app usage metric can be used to estimate the reduction in the expected number of onward transmissions per infected individual, R(t), due to the direct impact of exposure notification influencing the quarantine, testing and ultimately isolation behavior of infectious persons. Note that the reduction in R(t) that we estimate is independent from its absolute value at any point in time, an absolute value that we do not estimate here.

The relative reduction in R(t) could be ∼11.4% if 1) all infected persons carried their phones with them at the time transmission occurred, 2) all individuals who infect others test positive, and do so rapidly enough to notify those they infect in time to prevent tertiary cases, 3) the app detects all exposures that led to transmission, and 4) notifications eliminate forward disease transmission by sufficiently influencing the behavior of notified infected persons. In other words, there will be an 11.4% reduction in R(t) when exposure notifications are received by and trigger timely quarantine in 11.4% of infected individuals. The direct reduction in R(t) will be smaller, because of violations in the four assumptions, especially the fourth given reports of low quarantine compliance.^8, 9^ However, R(t) is also indirectly reduced when the far larger number of exposure events that do not lead to transmission also lead to behavior change, which either prevents infection in the notification recipient, or elicits first quarantine and then isolation after they have been infected by a different exposure within the same social network.^10^ Indeed, empirical estimates of the epidemiological impact of app usage are higher than expected from models of their direct effects.^1^

Unlike other recent evaluations^1, 11-13^, we have not integrated over the timecourse of case counts (Figure 1) to estimate how many cases were averted through this reduction in R(t). Doing so would require the assumption that both imposed and voluntary mitigation measures would have been the same with vs. without the app, an assumption not supported by data.^14^ The new usage metric we propose in the current work, with its interpretation in terms of relative reduction in R(t), is less dependent on the particulars of a situation, and thus more appropriate for comparing evaluations of apps deployed under various levels of disease prevalence. In particular, unlike estimates of the numbers of cases averted,^1, 11-13^ our metric does not increase with the duration or intensity of an outbreak, and thus is more appropriate for comparisons across time and place. Interestingly, holding adoption constant according to our usage metric, the value of this technology is higher with low infection prevalence than with high.^15^

Because persons who tested positive were asked whether they had entered verification codes prior to case investigation interviews, our results demonstrate that notification via the Covid Watch app was more rapid both than traditional contact tracing at the University of Arizona, and than possible digital exposure notification workflows where case investigators provide verification codes over the phone. Like other jurisdictions that have automated code distribution (e.g., ^11^), we saw a dramatic increase in the frequency with which persons testing positive received and entered verification codes, following the switch to automation. Our automated code delivery via an end-user test results portal was subsequently adopted by commercial test providers in Arizona.

Our installation rate of 46% among cases can be compared to the 28% installation rate among the general population of England and Wales, to which a 25% reduction in the size of the second wave has been attributed.^1^ While a 55% rate of code usage might seem low, we note that a similar 55% rate has been seen in Germany.^16^ A higher rate of 72% was achieved in England and Wales.^1^ Even when the workflow of code receipt and entry is intuitive and requires only minimal action by app users, many infected persons who have previously downloaded the app have done so in order to receive rather than to trigger notifications, or did not anticipate how they would feel upon testing positive. Using the app to notify others is strictly voluntary. Usage rates can be much lower with inferior code delivery workflows, as seen in our own study before September 9.

Other jurisdictions may not have suitable analytics for measuring code usage rate among app-using cases. Comparisons could still be made for the total number of codes claimed divided by the total number of cases (corresponding to ∼25% in our study) – this metric combines both app installation and code usage. One complication in making this comparison is that many U.S. States issue codes to all cases, including non-app users, via automated SMS.^17, 18^ In this workflow, the receipt of a code may prompt app download and code entry without generating any exposure notifications, and so to avoid inflating the usage statistic, the number of codes that led to uploading Temporary Exposure Keys for zero days would need to be subtracted from the total. In the U.S. state of Washington, this metric thus has an upper bound of 5.1% (averaged across 1.8% before the SMS system was implemented and 9.6% after).^11^ A press report for California^19^ suggests ∼27,000 code entries, however the exact timeframe of study was not provided in the press report making even a rough estimate of cases impossible. California has had >3.6 million reported cases as of the time of writing, with the main wave of cases, roughly half the total, occurring during the period that exposure notifications were active.^20^

To promote app installation by end-users, most U.S. States have relied on “push” notifications sent out by Apple and Google directly to smartphone users’ phones. These were not available at the time of this study, and are currently available only for Apple’s own Exposure Notification Express system and not for custom iOS apps.^21^ Another strategy to promote adoption is to use the same app to check in to venues (e.g. restaurants) as a privacy-preserving alternative to giving personal details;^1^ this is unavailable in jurisdictions that do not collect venue attendance for possible contact tracing purposes. Here we have shown that it is possible to achieve high adoption within a tightly interconnected community using a social influencer marketing strategy, without the advantage of push notifications or a multi-purpose app.

## Conclusions

Here we have proposed a new metric for assessing usage of a contact notification app within a tightly interconnected community that does its own testing and tracing. Usage on the University of Arizona campus is high enough to make it an impactful tool that complements and augments traditional contact tracing. While our study is limited to a specific population and time, it raises the possibility that other highly interconnected communities such as college campuses, large workplaces, tribal nations, and congregate living settings could benefit from targeted adoption campaigns even in the absence of statewide adoption and promotion. Further evaluation is needed to assess the extent of compliance with quarantine among infected persons receiving notification via such apps, which is a key determinant of the overall impact of exposure notification on reducing SARS COV-2 transmission.

## Data Availability

Aggregate data is available upon request.

## Acknowledgments

We thank Dr. Theresa Cullen for access to Pima County Health Department case investigation de-identified data, Janet McIllece for compiling the disparate data sources in a central location, and Amy Glicken for clarifications regarding university testing procedures.

## Research ethics

Data were collected under public health surveillance guidelines as part of an evaluation of the system. A request for aggregate data was made by the evaluation team to the Pima County Health Department and University of Arizona privacy officers in consultation with the University of Arizona IRB and was deemed not human subjects research. Data on positive tests were publicly available on the University of Arizona dashboard.

## Data availability

Aggregated data are available upon request.

